# An integrative examination of psychological distress and its nutritional and visual correlates among young adults in Ghana

**DOI:** 10.1101/2025.08.01.25332578

**Authors:** Isaiah Osei Duah Junior, Wendy Ofori Asare, Elizabeth J. Johnson, Charllote Boateng, Hubert Osei Acheampong, David Ben Kumah, Kwadwo Owusu Akuffo

**Affiliations:** Department of Optometry and Visual Science, College of Science, Kwame Nkrumah University of Science and Technology, Kumasi, Ghana; Department of Biological Sciences, College of Science, Purdue University, West Lafayette, IN 47906, United States of America; Friedman School of Nutrition Science and Policy, Tufts University, Boston, MA, United States of America; School of Public Health, College of Health and Allied Sciences, Kwame Nkrumah University of Science and Technology, Kumasi, Ghana; School of Public Health, University of Memphis, Memphis, TN 38152, United States of America; Department of Anatomy and Cell Biology, School of Medicine, Wayne State, Detroit, MI 48201, United States of America

**Keywords:** lutein, zeaxanthin, vision–related quality of life, risky behaviors, substance use, mental health disorders, psychological distress, young adults

## Abstract

Psychological distress is a key precursor to suicidal ideation during emerging adulthood—a period characterized by increasing independence and responsibility. Although young adults are particularly vulnerable, the burden and determinants of psychological distress among this group, especially vicenarians, remain poorly understood. This study utilized integrative approach to investigate psychological distress and its nutritional and visual correlates among 301 young adults in Ghana. Biographical, health, and physical activity data were collected using structured questionnaires. Visual function was assessed objectively using LogMAR Early Treatment Diabetic Retinopathy Study (ETDRS) charts for best corrected visual acuity (BCVA), Pelli-Robson charts for contrast sensitivity, and subjectively using the National Eye Institute Visual Function Questionnaire-25 (NEI-VFQ-25). Anthropometric measurements followed standard protocols. A 3-day 24-hour dietary recall was used to estimate carotenoid intake. Neuropsychological function was assessed with a cognitive test battery, and psychological distress was measured using the Kessler Psychological Distress Scale (K10). Logistic regression analyses revealed that 23.26% of participants experienced psychological distress—14.6% mild, 3% moderate, and 5.6% severe. Psychologically distressed individuals reported lower intake of lutein and zeaxanthin and were less likely to use corrective eyewear. Alcohol consumption increased the odds of distress, whereas better self-reported visual function (NEI-VFQ-25) reduced it. These findings suggest that integrating nutrition, substance use counseling, and eye care into mental health services may enhance psychological wellbeing among young adults in Ghana.

## Introduction

Optimal mental health is vital for survival, effective functioning, and healthy social relationships. When mental health deteriorates, young adults may develop suicidal thoughts, posing a serious public health concern. [1, 2]. Emerging adulthood is a fast-paced transitional phase in which vicenarians take on responsibilities, make independent decisions, and pursue financial independence [3]. This period often involves intimate relationships [4] and demanding academic pursuits [5] which can limit physical activity [6]. Increased independence may also encourage sedentary lifestyles and risky behaviors such as poor diet, alcohol use, and smoking [7]. Combined, these factors can disrupt the balance between personal and academic life, weakening mental health resilience and increasing the risk of psychological distress [8].

Psychological distress—a mental health condition marked by emotional suffering, anxiety, depression, and nervousness—is a major precursor to suicidal ideation [9]. In young adults, undiagnosed mild stress can progress into chronic distress, heightening the risk of mild cognitive impairment. Higher levels of psychological distress are also linked to reduced cognitive function and accelerated cognitive decline [10]. Evidence suggests that optimizing cognitive health through dietary carotenoid intake may help protect against psychological distress [11–13].

Dietary carotenoids like lutein and zeaxanthin have been linked to enhanced cognitive function across the lifespan [14]. Predominantly found in fruits, vegetables, sweet corn, and eggs, these carotenoids offer antioxidant, anti-inflammatory, neuroprotective, and light-filtering benefits that support both cognitive and visual health [14–17]. However, studies indicate that young adults often consume inadequate amounts of fruits and vegetables, suggesting low intake of lutein and zeaxanthin [18–20]. Since low levels of these carotenoids are associated with diminished cognitive function and heightened psychological stress [12, 15, 21], it is plausible that this population experiences elevated psychological distress [12].

Several studies have reported an inverse relationship between visual function and psychological distress [22–24], with additional findings showing that individuals with self-reported visual difficulties often experience higher levels of psychological distress [25–27]. Emerging adulthood is associated with intense academic demands that strain the visual system, increasing the risk of asthenopic symptoms [28] and induced refractive errors [29]. Unrecognized and untreated visual issues can contribute to psychological distress in this at-risk population [27, 30].

High-risk behaviors, particularly alcohol use, are common during emerging adulthood [31–33], often driven by social pressure, experimentation, stress coping, and the need for peer acceptance [34–36]. Alcohol consumption, especially during special occasions, can acutely impair memory, judgment, decision-making, attention, and coordination, potentially leading to psychological distress [37]. Combined disruptions in cognitive function and mental stability may increase involvement in social vices, posing broader risks to society.

Psychological distress exists on a continuum from mild to severe, and failure to identify and address it early can lead to suicide. While young adults are particularly vulnerable, the burden and contributing factors of psychological distress among this group in Ghana remain underexplored. This study assessed the psychological distress and associated nutritional and visual correlates using a multimodal approach combining self-reports and clinician assessments. Findings revealed a high burden, with one in five participants affected. Psychological distress was significantly influenced by diet, alcohol consumption, and visual function. These results underscore the need for multidisciplinary interventions to address psychological distress in this population. Future longitudinal studies should explore the potential of lutein and zeaxanthin supplementation, vision correction, and reduced alcohol intake to mitigate psychological distress in emerging adults.

## Materials and methods

### Methods

#### Study design, population, and area

This study utilized data collected between July 12 and August 2, 2021, as part of a larger research project evaluating macular pigment optical density (MPOD) in a cohort of healthy young adults in Ghana [20]. In brief, systematic random sampling combined with proportionate to size was used to enroll 301 healthy volunteers (defined as subjects with no ocular lesions and no neurological diseases who could tolerate the MPOD test). A hierarchical methodology was used, with biographical data, health status, lifestyle, physical activity, diet, anthropometry, visual function, neuropsychological performance and psychological distress collected either by structured questionnaire or standardized clinical assessment, as appropriate, and where necessary, by an optometric resident and well-trained clinical research assistants [20].

#### Measures

Biographical variables in our analysis include age, sex, ethnicity; health status variables include systolic and diastolic blood pressure and medication use; lifestyle variables include smoking history, exposure to passive smoking, alcohol consumption, light exposure, protective eyewear and physical activity measures; type and duration of physical activity.

#### Anthropometric measures

Bioelectric impedance analysis using the Seca mBCA525 portable device was used to assess fat mass index (FMI) and skeletal muscle mass index (SMMI), together with estimation of body weight and height to calculate body mass index (BMI) [54]. BMI categorized participants into underweight (<18.5), standard (18.5–24.9), overweight (25.0–29.9), and obese (≥30) [55].

#### Dietary measures: lutein and zeaxanthin intake

Twenty-four-hour dietary recall of a three-day dietary pattern (two weekdays and one weekend) guided by food modules and household handles assessed as previously described to investigate carotenoid intake [20, 56]. Lutein and zeaxanthin intakes were estimated with a validated dietary questionnaire [20].

#### Visual function measures

Visual function data were collected by optometric residents. Briefly, monocular visual acuity was measured using the logarithm of the minimum angle of resolution (LogMAR) of the Early Treatment Diabetic Retinopathy Study (ETDRS) at four meters and contrast sensitivity function was assessed using the Pelli-Robson chart at one metre [20]. In vivo macular pigment optical density, a surrogate measure of retinal and brain carotenoids, was measured at half and one degree of retinal eccentricity using adapted heterochromatic flicker photometry [20]. The MPOD utilizes iso-illuminance matching of the peripheral, non-absorbed green flicker and the central, maximally absorbed, blue, non-flicker circular stimulus (log I_central_/I_peripheral_) [57]. MPOD data were quantified from the eyes with the best corrected visual acuity (BCVA) and/or the dominant eyes using the Miles technique for equal monocular BCVA [58]. Further, subjective visual function, which measures the impact of visual impairment on various aspects of a person’s life, including daily activities, social functioning and emotional well-being, was assessed using the National Eye Institute Visual Function Questionnaire-25 (NEI VFQ-25). The NEI VFQ-25 scale ranges from 0 to 100, with higher scores indicating better visual functioning [59].

#### Neuropsychological function measures

Global cognition (attention, orientation, memory, and spatial recognition) were assessed with a 30-point dementia and/or Alzheimer’s disease screening tool, the Mini-Mental State Examination (MMSE) [20, 60], phonetic verbal fluency was investigated with the “FAS Test” as a function of language, memory, and executive functions; and Animal fluency tests were used to interrogate the semantic memory and verbal fluency [20, 61].

#### Psychological distress measures

The primary dependent variable was psychological distress, measured by the Kessler-10 Psychological Distress Scale (K-10) [62]. The scale consists of 10 items (e.g., ‘In the past 4 weeks, how often have you felt so nervous that nothing could calm you down?’), with each item having a five-point response scale: ‘always’ (5), ‘most of the time’ (5), ‘most of the time’ (4), ‘sometimes’ (3), ‘some of the time’ (2), and ‘never’. The scale ranges from 10 to 50, with a score < 20 indicating low/minimal distress, 20-24 indicating mild distress, 25-29 indicating moderate distress, and a score ≥ 30 indicating severe distress [20]. The Kessler-10 scale is related to the Composite International Diagnostic Interview (CIDI), a standard tool for the assessment of mental disorders, and remains a robust tool for the assessment of psychological distress[62, 63].

#### Ethical approval

The study was approved by the Institutional Review Board of the Kwame Nkrumah University of Science and Technology, Committee on Human Research Publication and Ethics at the School of Medicine and Dentistry (reference number: CHRPE/AP/198/21). Written informed consent was obtained voluntarily from all participants before enrolment, and all procedures used in the study conformed to the tenets of the Declaration of Helsinki.

#### Data analysis

Statistical Package and Service Solution version 25, which is compatible with Windows 10, was used to analyze the data. Shapiro-Wilk statistics were used to test the normality of the data. Where appropriate, independent t-test, for continuous normally distributed variables, or Mann-Whitney

for continuous skewed variables, and chi-squared analysis for categorical variables, were used to test the differences in estimates of psychological distress and all explanatory variable measures: biographical, health, physical activity, diet, musculoskeletal, visual, neuropsychological function in the study and their corresponding Hedge effect size computed. To examine the association between each explanatory variables on psychological distress a bivariate logistic regression analysis was performed at a significance of p≤0.05. The statistically significant variables in the bivariate model were included in the multiple logistic regression analysis and their interactive association with psychological distress ascertained at an arbitrary significance of p≤0.05. Odds ratios (OR) with 95% confidence intervals were calculated. The binary outcome was coded by dichotomizing the Kessler-10 scale into two categories with a total score < 20 for no psychological distress (coded 0) and ≥ 20 for mild to severe psychological distress (coded 1) [25, 64].

## Results

### Psychological distress was unrelated to biographical factors, health, or physical activity, but linked to lifestyle factors in participants

Table 1 summarizes the biographical, health, lifestyle, and physical activity characteristics of the participants. The overall prevalence of psychological distress was 23.26%, comprising mild (14.6%), moderate (3%), and severe (5.6%) cases. No significant differences were observed in age, sex, ethnicity, blood pressure, or medication use between participants with and without psychological distress (p > 0.05). None of the participants reported a history of smoking, and exposure to passive smoking or light did not significantly differ between groups (p > 0.05). However, a near-significant difference was noted in alcohol consumption (p = 0.051), and a statistically significant difference was found in eyeglass use (p = 0.030), with less eyeglass use among those reporting psychological distress. No significant association was observed between psychological distress and the type or duration of physical activity (p > 0.05).

**Table 1:**
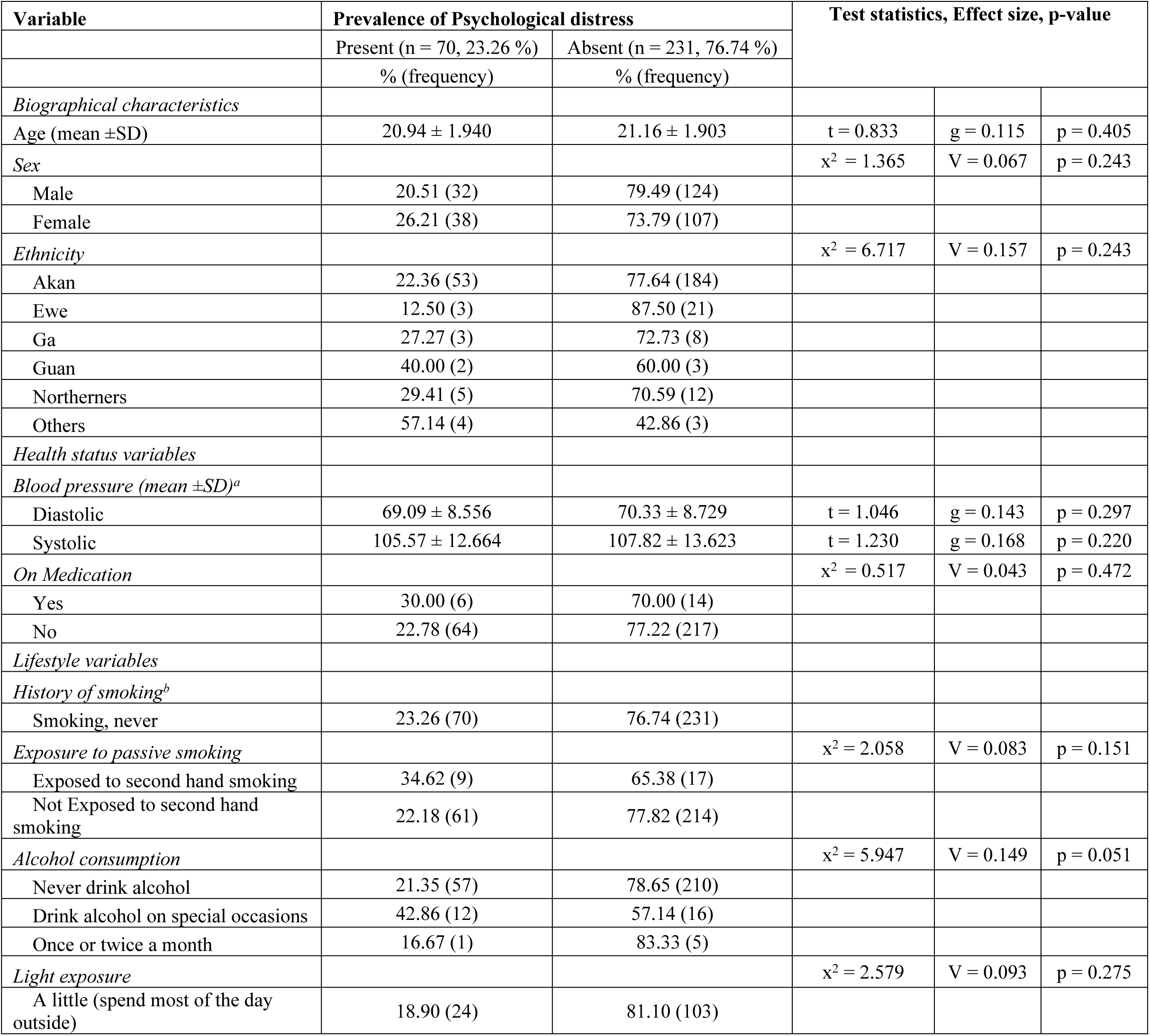
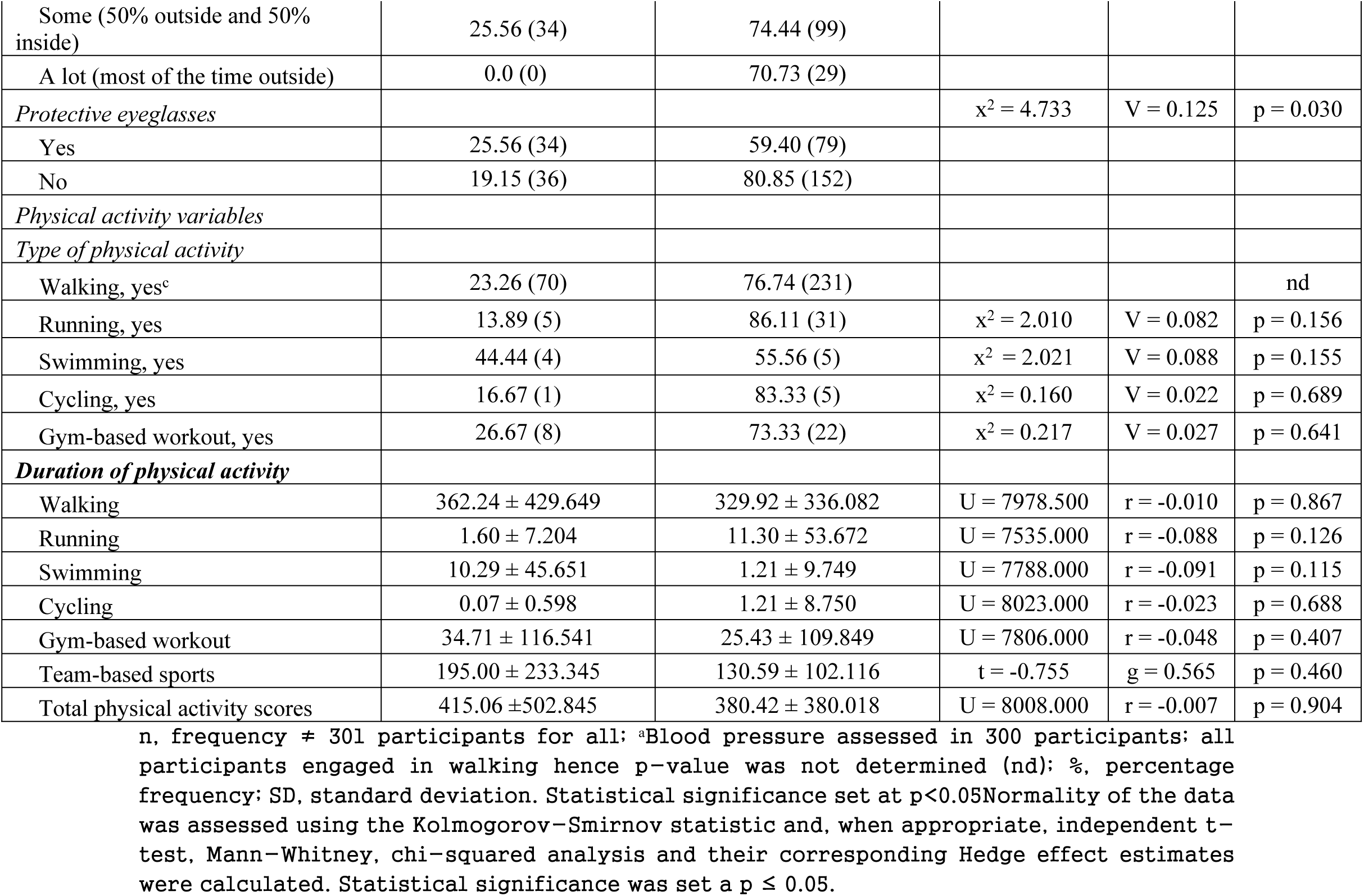
Biographical, health status, lifestyle, and physical activity measures on psychological distress.

### Dietary carotenoids and self-reported visual function, but not objective vision, musculoskeletal, or cognitive function, are related to psychological distress

**Table 2** presents the differences in psychological distress in relation to dietary intake of lutein and zeaxanthin, body composition, visual performance, and cognitive measures. Participants with low intake of lutein (p = 0.014), zeaxanthin (p = 0.026), or both (p = 0.022) were significantly more likely to report psychological distress. In contrast, the use of vitamins or supplements was not associated with psychological distress (p = 0.110). No significant differences were observed across body composition metrics—including fat mass index, skeletal muscle mass index, body mass index, and waist circumference (p > 0.05). While objective visual measures (visual acuity and contrast sensitivity) were comparable between groups, those with psychological distress had significantly lower subjective visual function scores (p = 0.001). Although not statistically significant (p > 0.05), macular pigment optical density tended to be lower in the psychologically distressed group. Additionally, no significant differences were observed in global cognitive performance across three neurocognitive platforms (p > 0.05).

**Table 2:**
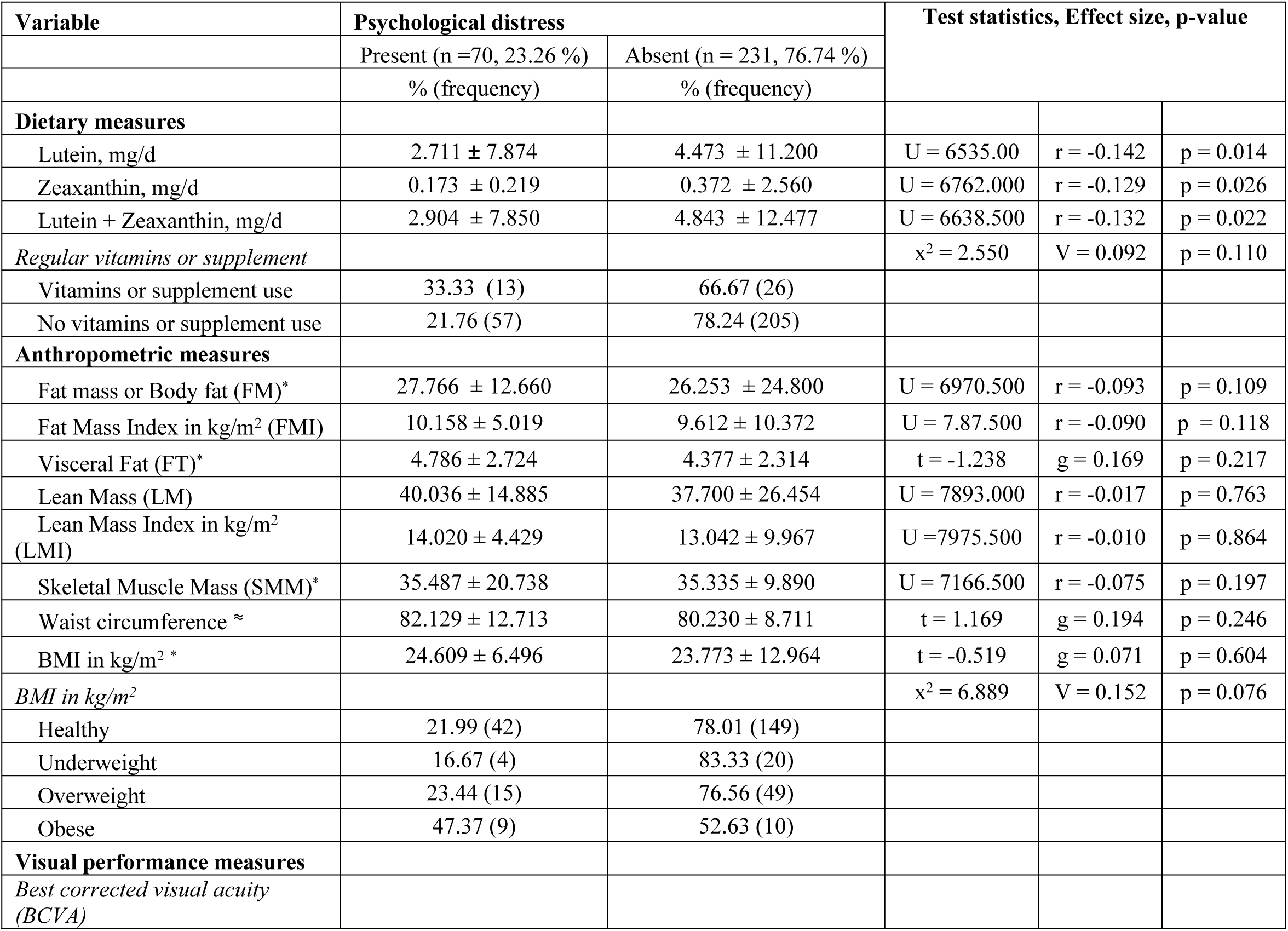
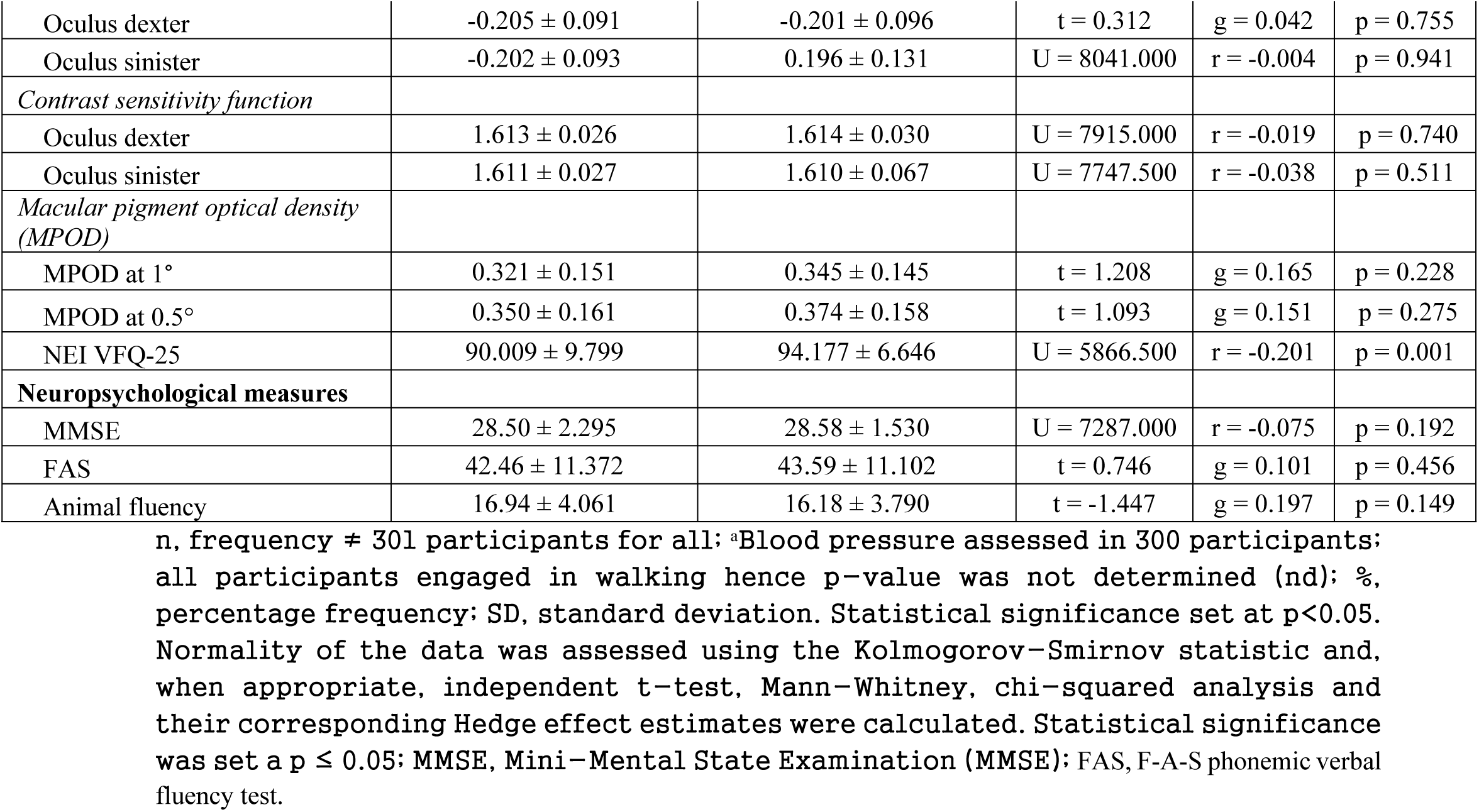
Dietary, musculoskeletal, visual function and neuropsychological function measures on psychological distress.

### Alcohol intake and subjective visual function counteract psychological distress

**Table 3** presents the bivariate and multiple regression analyses examining associations between psychological distress and various biographical, health, lifestyle, physical activity, dietary, body composition, visual, and cognitive measures. After adjusting for covariates, alcohol consumption on special occasions was significantly associated with increased odds of psychological distress (Adjusted Odds Ratio [AOR]: 3.032, 95% CI: 1.259–7.300, *p* = 0.013). In contrast, higher self- reported visual function was significantly associated with a lower likelihood of experiencing psychological distress (AOR: 0.944, 95% CI: 0.910–0.978, *p* = 0.002).

**Table 3:** Bivariate and multiple logistic regression analysis of factors associated to psychological distress.

| Variable | Bivariate regression |  |  | Multivariate regression |  |  |
| --- | --- | --- | --- | --- | --- | --- |
|  | COR | 95% CI (Lower, Upper) | p-value | AOR | 95% CI (Lower, Upper) | p-value |
| Biographical measures |  |  |  |  |  |  |
| Age, years | 0.941 | 0.817, 1.085 | 0.404 |  |  |  |
| *Sex* |  |  |  |  |  |  |
| Male | Ref |  |  |  |  |  |
| Female | 1.376 | 0.805, 2.354 | 0.244 |  |  |  |
| *Ethnicity* |  |  |  |  |  |  |
| Akan | Ref |  |  |  |  |  |
| Ewe | 0.496 | 0.142, 1.727 | 0.271 |  |  |  |
| Ga | 1.302 | 0.334, 5.081 | 0.704 |  |  |  |
| Guan | 2.314 | 0.377, 14.215 | 0.365 |  |  |  |
| Northerners | 1.447 | 0.488, 4.290 | 0.506 |  |  |  |
| Others | 4.629 | 1.005, 21.330 | 0.049 | 1.983 | 0.357, 11.009 | 0.434 |
| **Health status measures** |  |  |  |  |  |  |
| *Blood pressure , mm Hg* |  |  |  |  |  |  |
| Diastolic | 0.983 | 0.953, 1.015 | 0.296 |  |  |  |
| Systolic | 0.987 | 0.967, 1.008 | 0.220 |  |  |  |
| *Medication Use* |  |  |  |  |  |  |
| Yes | 1.453 | 0.537, 3.935 | 0.462 |  |  |  |
| No | Ref |  |  |  |  |  |
| **Lifestyle measures** |  |  |  |  |  |  |
| *Exposure to secondhand smoke* |  |  |  |  |  |  |
| Exposed to secondhand smoke | 1.857 | 0.789, 4.374 | 0.157 |  |  |  |
| Not Exposed to secondhand smoking | Ref |  |  |  |  |  |
| *Alcohol consumption* |  |  |  |  |  |  |
| No alcohol | Ref |  |  |  |  |  |
| Alcohol on occasion | 2.763 | 1.237, 6.172 | 0.013 | 3.032 | 1.259, 7.300 | 0.013** |
| Alcohol use once or twice a month | 0.737 | 0.084, 6.433 | 0.782 |  |  |  |
| *Light exposure* |  |  |  |  |  |  |
| Spends <50% of the day outside | Ref |  |  |  |  |  |
| 50% outside and 50% inside | 1.474 | 0.816, 2.661 | 0.198 |  |  |  |
| Spends >50% of the time outside | 1.776 | 0.793, 3.977 | 0.163 |  |  |  |
| *Protective eyewear* |  |  |  |  |  |  |
| Yes | 1.817 | 1.057, 3.124 | 0.031 | 1.436 | 0.790, 2.611 | 0.236 |
| No | Ref |  |  |  |  |  |
| **Physical activity measures** |  |  |  |  |  |  |
| *Routine physical activity* |  |  |  |  |  |  |
| Running, yes | 0.496 | 0.185, 1.329 | 0.163 |  |  |  |
| Swimming, yes | 2.739 | 0.715, 10.494 | 0.141 |  |  |  |
| Cycling, yes | 0.655 | 0.075, 5.702 | 0.702 |  |  |  |
| Gym-based workout, yes | 1.226 | 0.520, 2.889 | 0.642 |  |  |  |
| *Duration of physical activity* |  |  |  |  |  |  |
| Walking | 1.000 | 1.000, 1.001 | 0.511 |  |  |  |
| Running | 0.977 | 0.949, 1.006 | 0.115 |  |  |  |
| Swimming | 1.015 | 1.001, 1.029 | 0.035 | 1.012 | 0.999, 1.026 | 0.073 |
| Cycling | 0.947 | 0.815, 1.100 | 0.475 |  |  |  |
| Gym-based workout | 1.001 | 0.998, 1.003 | 0.543 |  |  |  |
| Team-based sports | 1.005 | 0.993, 1.016 | 0.449 |  |  |  |
| Total physical activity scores | 1.000 | 1.000, 1.001 | 0.537 |  |  |  |
| **Dietary measures** |  |  |  |  |  |  |
| Lutein, mg/d | 0.981 | 0.950, 1.012 | 0.228 |  |  |  |
| Zeaxanthin, mg/d | 0.412 | 0.091, 1.863 | 0.412 |  |  |  |
| Lutein + Zeaxanthin, mg/d | 0.981 | 0.951, 1.012 | 0.227 |  |  |  |
| *Regular vitamins or supplement* |  |  |  |  |  |  |
| Vitamins or supplement use | 1.798 | 0.869, 3.722 | 0.114 |  |  |  |
| No vitamins or supplement | Ref |  |  |  |  |  |
| **Anthropometric measures** |  |  |  |  |  |  |
| Fat Mass or Body Fat (FM) | 1.003 | 0.992, 1.013 | 0.630 |  |  |  |
| Fat Mass Index in kg/m<sup>2</sup> (FMI) | 1.005 | 0.980, 1.032 | 0.675 |  |  |  |
| Visceral Fat (FT) | 1.070 | 0.961, 1.192 | 0.217 |  |  |  |
| Lean Mass (LM) | 1.006 | 0.990, 1.021 | 0.483 |  |  |  |
| Lean Mass Index in kg/m<sup>2</sup> (LMI) | 1.024 | 0.967, 1.084 | 0.425 |  |  |  |
| Muscle Mass (MM) | 1.001 | 0.981, 1.021 | 0.933 |  |  |  |
| Waist circumference | 1.020 | 0.992, 1.048 | 0.157 |  |  |  |
| *BMI in kg/m<sup>2</sup>* |  |  |  |  |  |  |
| Healthy | 1.409 | 0.457, 4.349 | 0.551 |  |  |  |
| Underweight | Ref |  |  |  |  |  |
| Overweight | 1.531 | 0.452, 5.181 | 0.494 |  |  |  |
| Obese | 4.500 | 1.108, 18.271 | 0.035 | 2.777 | 0.603, 12.781 | 0.190 |
| **Visual performance measures** |  |  |  |  |  |  |
| *Best corrected visual acuity (BCVA)* |  |  |  |  |  |  |
| Oculus dexter | 0.634 | 0.037, 11.008 | 0.755 |  |  |  |
| Oculus sinister | 0.676 | 0.069, 6.667 | 0.737 |  |  |  |
| *Contrast sensitivity function* |  |  |  |  |  |  |
| Oculus dexter | 0.402 | 0.000, 3522.198 | 0.844 |  |  |  |
| Oculus sinister | 1.680 | 0.012, 240.128 | 0.838 |  |  |  |
| *Macular pigment optical density (MPOD)* |  |  |  |  |  |  |
| MPOD at 1° | 0.321 | 0.051, 2.033 | 0.228 |  |  |  |
| MPOD at 0.5° | 0.384 | 0.069, 2.139 | 0.275 |  |  |  |
| VFQ-25 | 0.940 | 0.910, 0.971 | 0.000 | 0.944 | 0.910, 0.978 | 0.002** |
| **Neuropsychological function** |  |  |  |  |  |  |
| MMSE | 0.974 | 0.838, 1.133 | 0.735 |  |  |  |
| FAS | 0.991 | 0.967, 1.015 | 0.455 |  |  |  |
| Animal fluency | 1.052 | 0.982, 1.128 | 0.149 |  |  |  |

Dependent Variable, Prevalence of Psychological Distress. OR, Odds ratio; CI, Confidence Interval; AOR, Adjusted Odds Ratio; VFQ-25, The National Eye Institute 25-Item Visual Function Questionnaire; MMSE, Mini-Mental State Examination; FAS, F-A-S phonemic verbal fluency test. Bivariate logistic regression $\leq 0.05$ considered for inclusion in the multivariate model; Statistical significance set at $p \leq 0.05$ .

## Discussion

This novel study aimed to determine the prevalence of psychological distress and to identify high-risk behaviors and modifiable factors associated with it among young adults in Ghana. The overall prevalence of psychological distress was 23.26%, indicating that nearly one in five participants were affected. Those experiencing psychological distress had significantly lower intakes of lutein and zeaxanthin and were less likely to use eyeglasses. After statistical adjustment, alcohol consumption emerged as a significant predictor, with individuals who drank alcohol being more likely to experience psychological distress. Conversely, higher self-reported visual function scores (NEI-VFQ-25) were significantly associated with a reduced likelihood of psychological distress.

Optimal mental health is essential for survival and the successful performance of daily activities [38]. In Ghana, young adults represent a significant portion of the population and are in a life stage marked by shifts in living conditions that influence lifestyle factors such as diet and physical activity. Reduced parental oversight during this period often increases susceptibility to high-risk behaviors, including substance abuse, alcohol consumption, and smoking [8]. Concurrently, this phase typically aligns with higher education, which brings intense academic demands that may strain the visual system and impact eye health [8]. Although the burden of psychological distress in this population has not been extensively studied, anecdotal reports indicate rising suicide rates, particularly among university students in Ghana—a key subgroup of emerging adults. Based on this context, we hypothesize that even among healthy young adults without clinically diagnosed chronic or terminal conditions, the prevalence of psychological distress is high and is linked to modifiable lifestyle and behavioral factors.

In this study, we found that one in every five participants experienced psychological distress, ranging from mild to severe. Our results are similar to studies from Africa [39, 40], Australia [41, 42], Asia [43–45], Europe [46, 47] and America [48] which report prevalence rates between 15% and 60% [39–48]. These results underscore the high vulnerability of young adults to psychological distress. The observed progression from mild to moderate symptoms highlights the urgent need for early screening to identify at-risk individuals and for rehabilitation services to provide effective coping strategies—measures that may help prevent the escalation of symptoms and reduce the risk of suicidal ideation.

We observed lower intakes of lutein and zeaxanthin among participants experiencing psychological distress. These xanthophyll carotenoids, predominantly found in fruits and vegetables, are selectively deposited in the retina and brain, where they play critical roles in visual and cognitive function [14, 15, 20]. Deficiencies in lutein and zeaxanthin have been linked to impaired visual and cognitive health, as well as increased vulnerability to psychological stress [12]. These findings support the need for dietary education initiatives that promote the consumption of lutein- and zeaxanthin-rich foods as a potential strategy to reduce the growing burden of psychological distress [12].

Unexpectedly, we observed that most participants without psychological distress wore eye glasses. Interestingly, Guan and colleagues have shown that myopic students who studied more intensively experienced a reduction in learning and physical anxiety after being provided with spectacles. [49]. Given the significant learning demands during young adulthood, we recommend vision screening and, if necessary, refractive error correction as protective interventions to reduce psychological distress in high-risk groups [49].

Alcohol intake was associated with a higher likelihood of psychological distress, consistent with findings by Balogun *et al*. [50] and Geisner *et al* [51]. who observed increased distress with heavier alcohol use [50, 51]. Together, reducing alcohol consumption and increasing abstinent days may help lower psychological distress in this population [51].

Participants with better self-reported vision, reflected by high NEI-VFQ-25 scores, were less likely to experience psychological distress. This aligns with previous studies that observed an increase in psychological distress in patients who self-reported vision problems [25, 26]. Since most participants without distress wore spectacles, we recommend eye care screening to detect early vision difficulties that may contribute to psychological distress in this population (Table 2).

## Strengths and limitation

Factors accounting for psychological distress among young adults in Ghana remains unclear. In this novel investigation, we have provided comprehensive evidence of an increased burden of psychological distress among young adults. Taken together, this demonstrates the intellectual merit of this study in generating prevalence data, which are essential to contribute to systematic reviews and inform future longitudinal studies. We have also identified high-risk behaviors and highlighted modifiable factors that may help to reduce the burden. These findings have significant wider implications that may revolutionize institutional and/or national intervention policies and pave the way for the development of coping strategies for this population at high risk of psychological distress. It is worth mentioning that our study has limitations. Assessment of lutein and zeaxanthin was based on dietary recall which may not reflect the true intake considering recall bias. However, this approach has shown to provide robust estimates that are consistent with plasma profiling of carotenoids and widely used in most clinical and/or epidemiological studies [52]. Intimate partner relationships are common among young adults and may also have an impact on mental health. Although our study did not investigate this, this may have obscured some findings, and future research to assess their influence on the increasing psychological distress in this population is a focus of future investigation [53].

## Conclusion

For every five young adults one was found to experience psychological distress, and most of these subjects had low intakes of xanthophyll carotenoids (lutein and zeaxanthin) and less use of eye wear. Alcohol consumption and increased self-reported vision were shown to have opposite associations, with the former increasing the likelihood of psychological distress and the latter being protective. The high burden calls for institutionalization of mental health screening programs to help detect early symptoms of psychological distress and timely provision of coping interventions and eye care services to identify and correct vision problems. Further, we recommend dietary education to help optimize lutein and zeaxanthin intake while decreasing alcohol consumption.

Collectively, this may ameliorate psychological distress and potentially contribute to reduce suicides among young adult population from Ghana.

## Data Availability

All relevant data and materials supporting the conclusion of this article is/are available within the manuscript and its supporting information files.

## Abbreviations

AOR: Adjusted odds ratio
BMI: Body mass index
BCVA: Best-corrected visual acuity
FMI: Fat mass index
CI: Confidence interval
LM: Lean mass
LMI: Lean Mass Index
SMM: Skeletal muscle mass
MPOD: Macular pigment optical density
MMSE: Mini-Mental State Examination (MMSE)
NEI-VFQ-25: National Eye Institute Visual Function Questionnaire-25 (NEI VFQ-25)
SD: Standard deviation
SMMI: Skeletal muscle mass index
OR: Odds ratio.

## Acknowledgements

The authors thank Professor Billy Wooten, Department of Psychology, Brown University, Providence, RI, USA, for the generous donation of a macular densitometer. We are indebted to Dr. Micheal Gyawu-Odame, Dr. Stephannie Obiri-Yeboah, and Dr. Stacy Ewurama Horthman of the Department of Optometry and Visual Science, College of Science, Kwame Nkrumah University of Science and Technology, Kumasi, Ghana, for their assistance with data collection. The authors are grateful to the research subjects for their willingness to participate.

## Author contributions

**I.O.D.J.:** conceptualization, methodology, project management, resources, experimental design, data curation, data validation, visualization, investigation, software, formal analysis, formatting, writing original draft, writing review and editing, correspondence. **W.O.A.:** methodology, project management, experimental design, data collection, data curation, investigation, software, formal analysis, resources, writing original draft, writing review and editing. **E.J.J.:** project management, technical assistance, data curation, resources, software, formal analysis, writing review and editing**. C.B:** resources, investigation, writing original draft, writing review and editing. **H.O.A:** resources, investigation, writing review and editing; **D.B.K**: methodology, investigation, resources, writing review and editing. **K.O.A.:** methodology, project management, resources, experimental design, data management, data validation, visualization, investigation, software, formal analysis, writing original draft, writing review and editing, and supervision.

## Declarations

### Consent for publication

Not applicable

